# Computational Analysis of Six Expression Studies Reveals miRNA-mRNA Interactions and 25 Consistently Disrupted Genes in Atopic Dermatitis

**DOI:** 10.1101/2022.06.04.22276002

**Authors:** Sarah Gao, Andrew Gao

## Abstract

Atopic dermatitis (AD), known as eczema, affects millions of people worldwide and is a chronic inflammatory skin disease. It is associated with risks of developing asthma, food allergies, and various other diseases related to the immune system. AD can also negatively affect the self-esteem of patients. Gene expression data could yield new insights into molecular mechanisms and pathways of AD, however, results often vary drastically between studies. In this study, expression data from five mRNA studies and one miRNA study were combined to identify differences between atopic dermatitis skin and unaffected, normal skin. Protein interaction network analysis and Panther analysis revealed that pathways related to leukocyte behavior, antimicrobial defense, metal sequestration, and type 1 interferon signaling were significantly affected in AD. In total, 25 genes, such as SERPINB4 and ST1007 were consistently identified to be disrupted across studies. Within the 25, 11 were underexpressed and 14 were overexpressed. Several genes implicated in skin cancers were among the 25. We also identified underexpressed 13 miRNAs, many of which regulate some of the 14 overexpressed genes. Gene FOXM1 was targeted by 6 underexpressed miRNAs and was on average overexpressed by 9.53 times in AD. Presumably, underexpression of miRNAs led to overexpression of their gene targets. The results of this research have implications for diagnostic tests and therapies for AD. It elucidates molecular mechanisms of AD with greater confidence than does a single study alone. Future steps include experiments regarding the role of SERPINB4, ST1007, neutrophil and leukocyte aggregation, and interferon signaling in AD. Additionally, the associations between AD and skin cancers should be further investigated.

## Introduction

Atopic dermatitis (AD), also called eczema, affects about ten to twenty percent of people in developed countries, such as the United States of America and France [1-2]. AD is known as a chronic inflammatory skin disease which means that it lasts for a long time and occurs when the immune system wrongly targets normal body tissues [3]. Treatments for AD amount to up to $3.8 billion dollars yearly in the United States alone [4].

AD is linked to higher risk for getting asthma, food allergies, and other immune system related diseases [5]. AD can also lower patient self esteem as it causes visible lesions and marks on the skin [6]. While AD has been researched before, scientists are still not entirely sure about how AD develops (pathogenesis) [7]. Therefore, research should be conducted to explore the pathways and mechanisms that cause, or are affected by, atopic dermatitis development. With this knowledge, new treatments and tests for AD can be developed and existing methods can be improved.

Current treatments for AD include using moisturizers and applying corticosteroids to the affected skin [8]. However, the effectiveness of the treatments vary greatly. For some people the medicines help a lot but for others the medicines don’t do anything.

Previously, microarrays have been used to measure gene and miRNA expression levels, often for disease research [9-12]. For example, microarrays are used to find differentially expressed genes (DEGs) between Parkinson’s patients and healthy control patients as well as differentially expressed miRNAs between lung cancer patients and healthy controls [13-14]. Microarrays are useful for discovering disease biomarkers and for learning more about changes in gene expression due to disease or some experimental variable [15]. While they have begun to be replaced by RNA-seq in many cases, existing microarray data still holds a lot of valuable information that can be analyzed.

Several teams have performed microarray analysis on AD patients and control patients in order to find differences in gene expression that could shine light on how AD develops. Some researchers have focused on gene expression while others have focused on miRNA expression [16-17]. MiRNAs are non-coding RNAs that have an important role in regulating gene expression [18]. They repress protein production by destabilizing the mRNA they are targeting. As miRNAs are being studied more, scientists are realizing that miRNA and gene expression are often intrinsically correlated [19]. Thus, research linking miRNA expression and gene expression in AD is needed. For example, a miRNA could change gene expression in many genes in a pathway important to AD.

Additionally, the existing studies conducted on AD gene/miRNA expression have small sample sizes, meaning that the results are more affected by random chance and individual differences in the patients from which skin samples were collected. Microarray experiments have been criticized in the past for often having errors and batch effects, among other things [20-22]. This means that a lot of the genes identified by one study are not able to be reproduced, or identified, by another group of researchers and sometimes the microarray data is flawed or biased.

In the present study, gene and miRNA expression data from multiple sources were combined to provide greater confidence and accuracy in the results as well as elucidate interactions between miRNAs and genes that can play a role in AD. Our approach emphasized biological relevance and statistical significance, often erring on the side of caution. For example, stringent minimum fold change and dataset overlap requirements were enforced.

## Methods

### GEO Dataset Search and Selection

The Gene Expression Omnibus (GEO) is an online database of freely available data from various experiments, usually with gene expression [23]. GEO was searched for datasets related to atopic dermatitis. Criteria for selecting a dataset included sample size, age of the data, source of sample (skin tissue only), and the type of AD examined. Six datasets were selected for further analysis: GSE32924, GSE31408, GSE36842, GSE16161, GSE5667, GSE6012 [24-29].

GSE31408 contained miRNA expression data while the rest contained mRNA expression data. Many of the studies were comprised of samples from several types of atopic dermatitis, such as chronic atopic dermatitis and nonlesional atopic dermatitis. We grouped all such types under the umbrella category of atopic dermatitis. While having different specific characteristics, different types of atopic dermatitis still are likely to share similarities and atopic dermatitis is known as a systemic disease. Thus the tradeoff of combining types of atopic dermatitis to gain greater sample size is justified.

### Data processing and statistical analysis

GEO2R is an online interactive tool that compares gene expression levels of groups of samples to identify genes that are differentially expressed between groups [30]. Following dataset selection, GEO2R was used to calculate differentially expressed genes between the AD samples and the control samples in each of the datasets. GEO2R automatically performed t-tests on the data. Expression intensity value distribution was normal in each dataset, showing no outliers. Thus, all samples were kept. Subtypes of atopic dermatitis were all included in the AD group, such as nonlesional and lesional. Depending on the study, differentially expressed messenger RNAs or differentially expressed microRNAs were found. The DEGs were saved into Excel sheets.

Differentially expressed genes with P values of greater than 0.05 were deleted. Genes were split into two lists, overexpressed and underexpressed (positive fold change and negative fold change, respectively). Next, we applied a minimum fold change threshold of 1.5 for overexpression and ⅔ for underexpression in an effort to enforce biological relevance standards and also reduce the number of genes for analysis (tens of thousands of genes passed the P value test).

### Identifying common genes across studies

After the relevant genes for each dataset were split into two lists, overexpressed and underexpressed, data were cleaned, removing duplicate genes and blank rows.

We combined the lists of overexpressed genes from each of the five mRNA datasets together. The same was performed for the underexpressed genes. Next, we used Python and Google Colab to calculate out how many times each gene was repeated in a list. This revealed how many times each gene was identified across datasets.

Only genes identified in at least four out of five datasets were kept and the rest were deleted (Common Data Ratio >= 0.8).

### STRING protein interaction network analysis

STRING is an online database search and network visualization tool of established and predicted protein interactions [31]. STRING was used to analyze relationships between the protein products of genes that were identified in previous steps. Overexpressed and underexpressed genes were analyzed separately. The cut off criteria for minimum confidence of genuine protein interaction was 0.4 which is accepted as medium confidence. Hub gene criteria was having at least a node degree of 6.

### Gene Ontology and pathway analysis

Gene Ontology (GO) is a popular computational biology tool that gives information on the functions of various gene products [32]. PantherDB, a tool by Gene Ontology, was applied to identify enriched biological functions [33].

### miRNA target prediction and analysis

The miRNA cutoff criteria were p-value <0.05 and fold change >1.5 or <0.66. The gene targets for the differentially expressed microRNAs identified in GSE31408 were predicted with the miRNet software. The gene targets were analyzed to find overlaps with the differentially expressed genes identified from the mRNA studies. It was assumed that under-expressed miRNAs would be associated with over-expressed target genes, and vice versa.

## Results

GSE32924, GSE6012, GSE36842, GSE16161, GSE5667, and GSE31408 were chosen for this study (Table 1). GSE31408 is a miRNA expression profiling study, while the other five are gene expression profiling studies. In total, there were 80 atopic dermatitis samples and 47 healthy control samples across all 5 gene expression datasets. There were 24 atopic dermatitis and 2 control samples in the miRNA dataset. Expression value distribution was checked for each dataset and all were normal, with no outliers.

**Table 1:** Descriptive information for each of the six Gene Expression Omnibus datasets analyzed in this study. Specifically, the columns indicate the number of AD samples, number of control samples, type of expression profiled, platform, author of study, year of study, and country of study.

| Dataset Accession | AD Samples | Control Samples | Type | Platform | Author | Year | Country |
| --- | --- | --- | --- | --- | --- | --- | --- |
| GSE32924 | 25 | 8 | Gene | GPL570 | Suárez-Fariñas | 2011 | United States |
| GSE6012 | 10 | 8 | Gene | GPL96 | Benson | 2006 | Sweden |
| GSE36842 | 24 | 15 | Gene | GPL570 | Suarez-Farinas | 2012 | United States |
| GSE16161 | 9 | 10 | Gene | GPL570 | Suarez-Farinas | 2009 | United States |
| GSE5667 | 12 | 5 | Gene | GPL96 | Plager | 2006 | United States |
| GSE31408 | 24 | 2 | miRNA | GPL14149 | Søkilde | 2011 | Sweden |

After applying the logFC (>1.5 or <⅔) and p-value criteria (p<0.05), the number of differentially expressed genes were greatly reduced (Table 2). In general, more downregulated genes were identified than upregulated genes with the exception of GSE5667. Genes were sorted by logFC value.

**Table 2:**
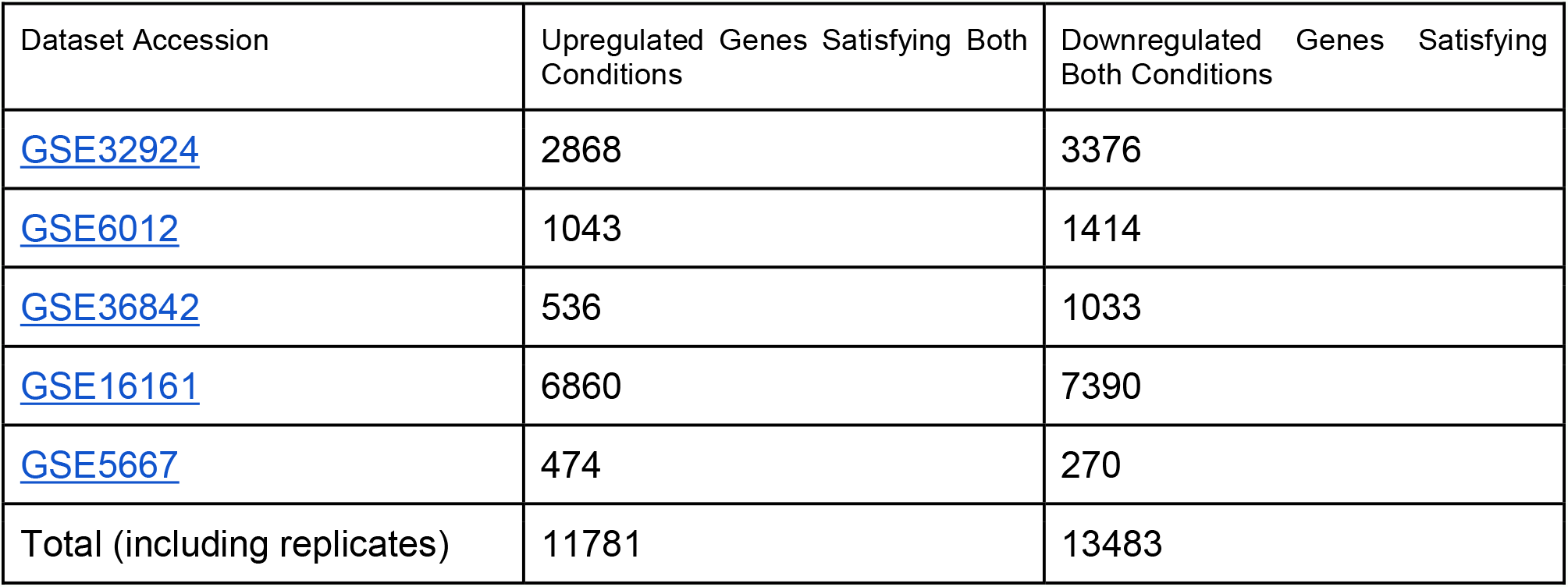
Number of genes identified as significantly upregulated or downregulated from each of the gene expression datasets. For instance, in dataset GSE32924, 2,868 upregulated genes passed the logFC and p-value criteria.

The five selected datasets contained a total of 11,781 upregulated genes and 13,483 downregulated genes satisfying both conditions (Table 3). There were a total of 5,874 unique upregulated genes across the five datasets: 371 of them were identified in only □datasets, 80 identified in □datasets, and 14 identified in all five datasets. There were a total of 6465 unique downregulated genes across the five datasets: 653 of them were identified in only □datasets, 103 identified in □datasets, and 11 identified in all five datasets. The majority of differentially expressed genes were only identified in one dataset out of five. Amongst the downregulated genes, 4,088 out of 6,565 total genes were only found in a single dataset (62.3%). Similarly, amongst upregulated genes, 3,802 out of 5,873 total genes were only found in a single dataset (64.7%). A total of only 25 genes were consistently upregulated or downregulated in all five studies (Table 4).

**Table 3:** Total unique genes. Also, the number of genes across all five datasets that were found in three, four, and five datasets. For instance, only 14 genes were identified as upregulated in all five datasets.

| Status | Total Unique Genes | Identified in only<br>□ datasets | Identified in only<br>□ datasets | Identified in 5/5<br>datasets |
| --- | --- | --- | --- | --- |
| Upregulated | 5874 | 371 | 80 | 14 |
| Downregulated | 6465 | 653 | 103 | 11 |

**Table 4:** 25 genes that appeared as upregulated (14) or downregulated (11) consistently in all five gene expression datasets. Genes are presented in alphabetical order.

| Gene name | Status |
| --- | --- |
| AKAP13 | Upregulated |
| DSC2 | Upregulated |
| DSG3 | Upregulated |
| FOXM1 | Upregulated |
| GART | Upregulated |
| H2AFX | Upregulated |
| IFI27 | Upregulated |
| LTB4R | Upregulated |
| MAP3K14 | Upregulated |
| MMP12 | Upregulated |
| NUP210 | Upregulated |
| S100A7 | Upregulated |
| SERPINB4 | Upregulated |
| TYMP | Upregulated |
| ALDH3A2 | Downregulated |
| BCHE | Downregulated |
| CHP2 | Downregulated |
| CST6 | Downregulated |
| IL37 | Downregulated |
| LEPROT | Downregulated |
| MSMB | Downregulated |
| PSORS1C2 | Downregulated |
| PTN | Downregulated |
| SCGB2A1 | Downregulated |
| TSPAN8 | Downregulated |

**Table 5:** Fold change for each of the 14 upregulated genes in each of the five datasets. The average fold change is also provided. The “miRNA regulated?” column shows whether the gene was identified as a target for one of the 13 underexpressed miRNAs, and if yes, how many miRNAs had it as a target. A value of 2 indicates that the gene was expressed twice as much in AD than in control skin.

| Gene | GSE32924 | GSE6012 | GSE36842 | GSE16161 | GSE5667 | Average FC (not log) | miRNA regulated? |
| --- | --- | --- | --- | --- | --- | --- | --- |
| SERPINB4 | 22.63 | 23.81 | 2.16 | 284.05 | 8.63 | 68.26 | no |
| S100A7 | 9.78 | 48.07 | 2.41 | 42.52 | 4.47 | 21.45 | no |
| MMP12 | 12.91 | 4.29 | 2.25 | 38.32 | 5.62 | 12.68 | no |
| FOXM1 | 3.92 | 2.89 | 1.55 | 37.01 | 2.28 | 9.53 | yes (6) |
| TYMP | 6.23 | 3.51 | 2.09 | 24.59 | 2.11 | 7.71 | yes (1) |
| DSC2 | 4.23 | 2.53 | 1.75 | 18.25 | 2.25 | 5.8 | yes (2) |
| IFI27 | 3.12 | 8.82 | 1.54 | 10.34 | 3.63 | 5.49 | yes (2) |
| NUP210 | 3.66 | 1.71 | 1.67 | 13.09 | 1.65 | 4.36 | yes (3) |
| MAP3K14 | 4.23 | 1.74 | 1.52 | 7.01 | 1.57 | 3.21 | yes (1) |
| H2AFX | 3.61 | 2.45 | 1.55 | 5.21 | 1.58 | 2.88 | yes (1) |
| GART | 2.28 | 1.55 | 1.59 | 4.5 | 1.92 | 2.37 | no |
| LTB4R | 2.66 | 1.06 | 1.61 | 5.03 | 1.51 | 2.37 | no |
| DSG3 | 1.78 | 2.45 | 1.22 | 4.06 | 1.57 | 2.22 | no |
| AKAP13 | 2.62 | 1.84 | 1.58 | 1.68 | 1.88 | 1.92 | yes (2) |

**Table 5:**
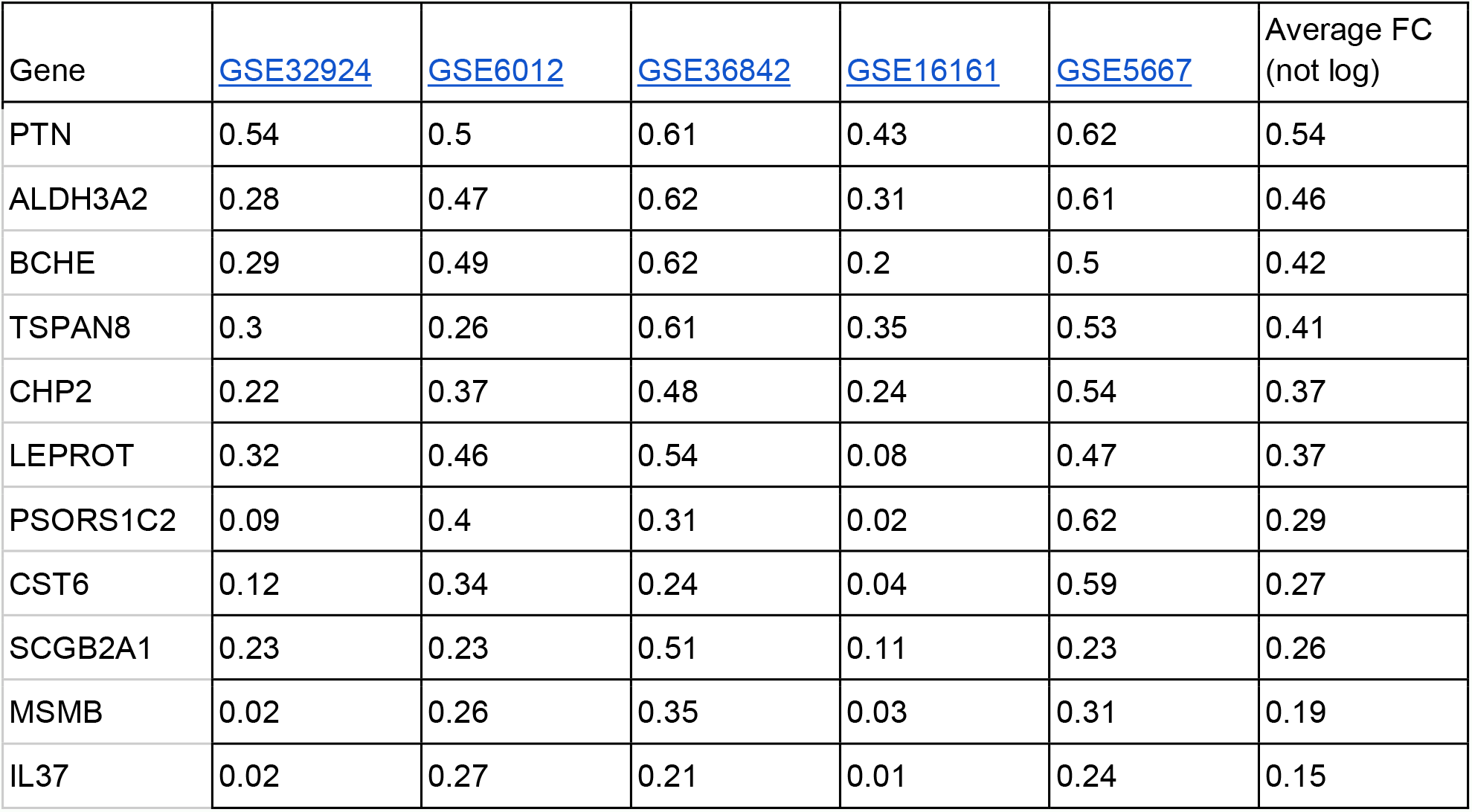
Fold change for each of the 11 downregulated genes in each of the five datasets. The average fold change is also provided. A value of 0.5 indicates that the gene was half as expressed in AD than control skin.

**Table 6:** Functional enrichment, p-value, and false discovery rates for the top six Gene Ontology biological process identified by Panther for downregulated genes.

| Downregulated Genes Panther Results |  |  |  |
| --- | --- | --- | --- |
| GO biological process complete | (fold enrichment) | p value | FDR (false discovery rate) |
| neutrophil aggregation (GO:0070488) | > 100 | 1.14E-04 | 1.74E-02 |
| sequestering of zinc ion (GO:0032119) | > 100 | 2.83E-04 | 3.54E-02 |
| leukocyte aggregation (GO:0070486) | 56.66 | 3.59E-05 | 7.42E-03 |
| sequestering of metal ion (GO:0051238) | 56.66 | 3.59E-05 | 7.32E-03 |
| leukocyte migration involved in inflammatory response (GO:0002523) | 40 | 8.86E-05 | 1.50E-02 |
| negative regulation of leukocyte mediated cytotoxicity (GO:0001911) | 34 | 1.36E-04 | 2.03E-02 |

**Table 7:** Functional enrichment, p-value, and false discovery rates for the top six Gene Ontology biological process identified by Panther for downregulated genes.

| Upregulated Genes Panther Results |
| --- |

| GO biological process complete | (fold Enrichment) | (raw P-value) | (FDR) |
| --- | --- | --- | --- |
| regulation of bone mineralization (GO:0030500) | 14.7 | 5.12E-06 | 9.04E-03 |
| regulation of biomineralization (GO:0110149) | 11.88 | 1.61E-05 | 1.83E-02 |
| regulation of biomineral tissue development (GO:0070167) | 11.88 | 1.61E-05 | 1.71E-02 |
| cardiac muscle tissue development (GO:0048738) | 8.25 | 2.98E-05 | 2.37E-02 |
| connective tissue development (GO:0061448) | 7.09 | 2.26E-05 | 2.00E-02 |
| ossification (GO:0001503) | 6.3 | 1.71E-05 | 1.70E-02 |

**Table 8:** 13 miRNAs identified as statistically relevant according to logFC and P-value.

| P.Value | logFC | miRNA_ID_LIST |
| --- | --- | --- |
| 0.00133333 | -1.89 | SNORD65 |
| 0.00076996 | -1.12 | hsa-miR-26a |
| 0.00017998 | -1.06 | hsa-let-7a |
| 0.0013918 | -1.04 | hsa-miR-143 |
| 0.00057997 | -9.92E-01 | hsa-let-7b |
| 0.00016403 | -9.54E-01 | hsa-miR-125b |
| 0.00204407 | -8.77E-01 | SNORD6 |
| 0.02253837 | -8.50E-01 | hsa-miR-24 |
| 0.00000262 | -7.12E-01 | hsa-miR-516a-3p,hsa-miR-516b* |
| 0.01926674 | -6.93E-01 | SNORD2 |
| 0.00007234 | -6.29E-01 | SNORD38B |
| 0.02638829 | -6.23E-01 | SNORD10 |
| 0.00072411 | -5.90E-01 | SNORD13 |

**Table 9:**
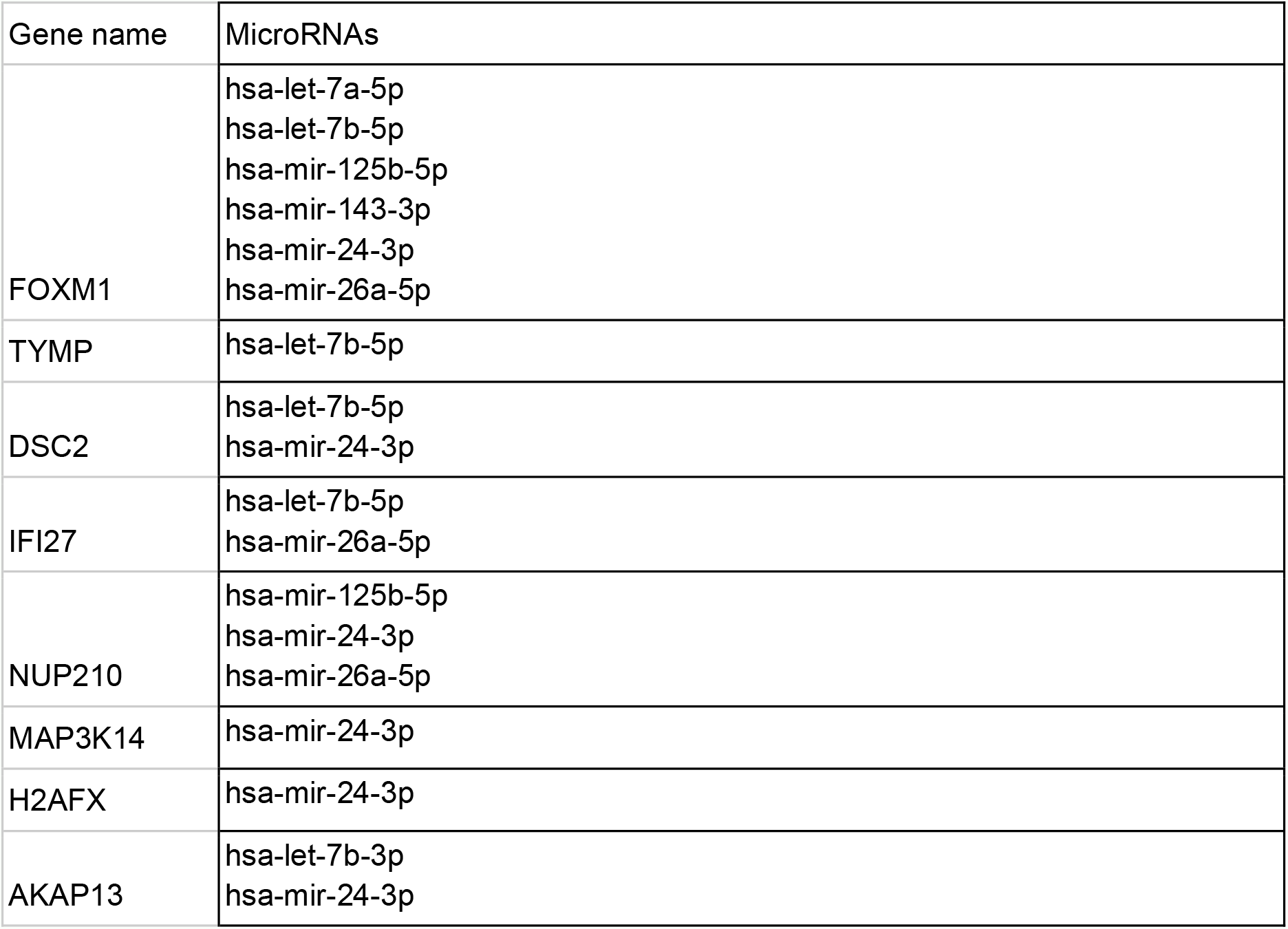
Upregulated genes identified as gene targets for at least one of the 13 downregulated microRNAs. FOXM1 was a gene target for six microRNAs.

| Gene name | MicroRNAs |
| --- | --- |
| FOXM1 | hsa-let-7a-5p<br>hsa-let-7b-5p<br>hsa-mir-125b-5p<br>hsa-mir-143-3p<br>hsa-mir-24-3p<br>hsa-mir-26a-5p |
| TYMP | hsa-let-7b-5p |
| DSC2 | hsa-let-7b-5p<br>hsa-mir-24-3p |
| IFI27 | hsa-let-7b-5p<br>hsa-mir-26a-5p |
| NUP210 | hsa-mir-125b-5p<br>hsa-mir-24-3p<br>hsa-mir-26a-5p |
| MAP3K14 | hsa-mir-24-3p |
| H2AFX | hsa-mir-24-3p |
| AKAP13 | hsa-let-7b-3p<br>hsa-mir-24-3p |

### Notable Genes

SERPINB4 appeared in all five datasets, and had the highest fold change out of all upregulated genes in four datasets, 68.26. SERPINB4 is in the SERPIN family of serine protease inhibitors [34]. SERPINB4 is a protein coding gene and it acts as an immune response gene. S100A7 was also found in all five datasets and regularly highly differentially expressed, with an average fold change of 21.45. Like SERPINB4, it was often among the top most upregulated genes in each dataset. S100A7 is S100 calcium-binding protein A7, or psoriasin [35]. It has antimicrobial activity and is reported to be produced by skin epithelial cells to ward off bacterial invaders. It also plays a role in the cell cycle. As its name psoriasin suggests, a study has found higher levels of S100A7 in psoriatic skin lesions. S100A9 was not identified as significantly differentially expressed in GSE5667 but appeared in the four other datasets. It belongs to the same family as S100A7, the S100 family, and also plays a role in the cell cycle [36]. S100A9 had an average fold change of 43.98, even higher than S100A7.

### STRING Network Analysis

Figure 1 shows all the different upregulated genes that appear in at least □ of the datasets, and the interactions between their protein products, visualized as edges between nodes. The diagram was generated using STRING DB and 92 out of 94 input genes were mapped (2 were not identified in the database search). 117 edges between gene nodes were mapped, with an expected value of 59 edges, demonstrating that the network of genes has significantly more interactions than would be expected of a random list. The average node degree was 2.54, meaning that on average each gene was linked to 2.54 others. Gene with node degree of at least 6 were: CXCR4, STAT1, SAMHD1, DDX58, IRF9, OAS1, HERC6, CDC20, MCM5, FOXM1, CDC6, TOP2A, S100A7, and KRT16. Notably, STAT1 had degree 12. STAT1 is a transcription factor involved in cytokine signaling, interferon response, and interleukin signaling [37].

**Figure 1:**
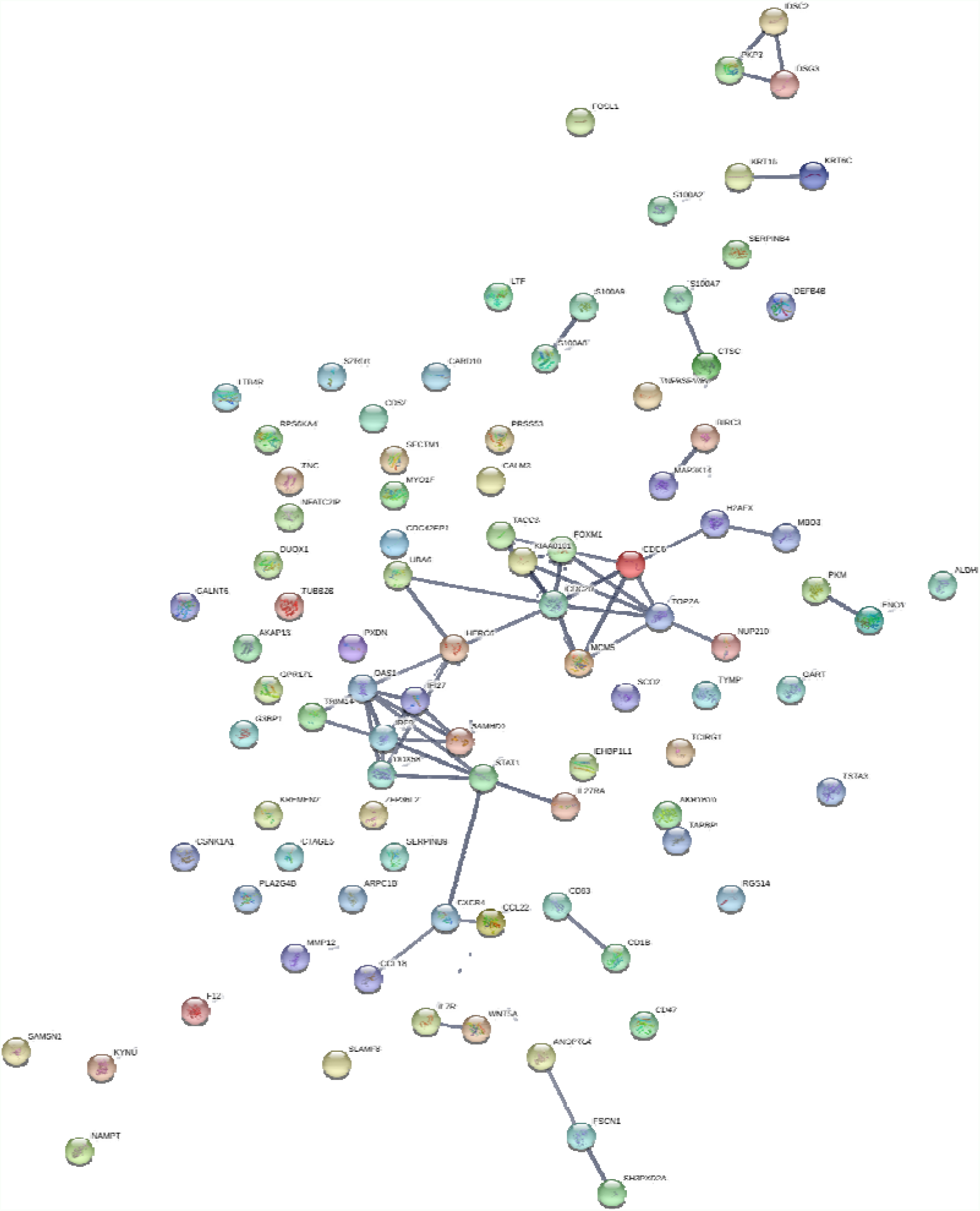
STRING protein interaction network for upregulated genes identified in at least four out of five datasets shows several highly interconnected clusters.

**Figure 2:**
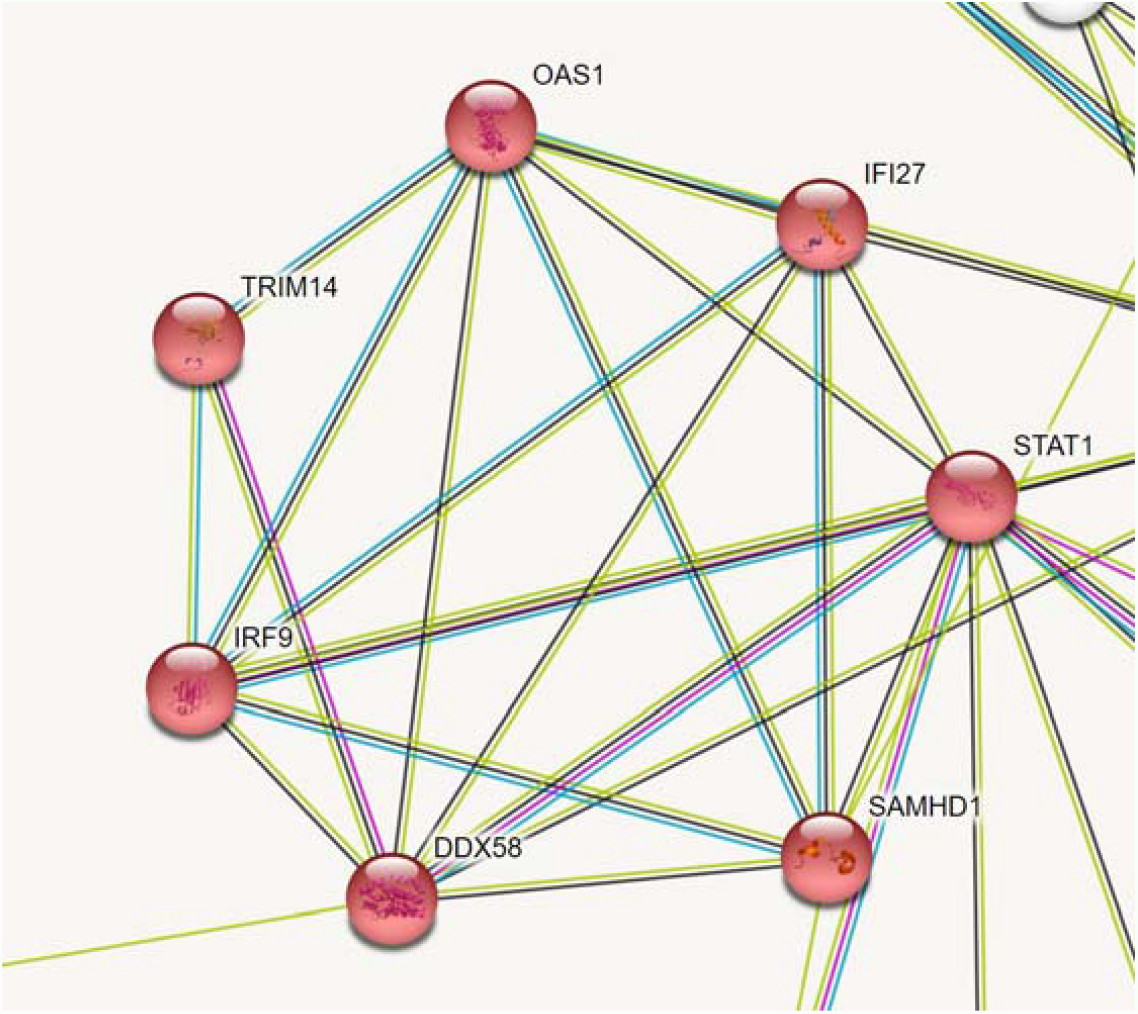
Interferon signaling genes cluster in STRING network

**Figure 3:**
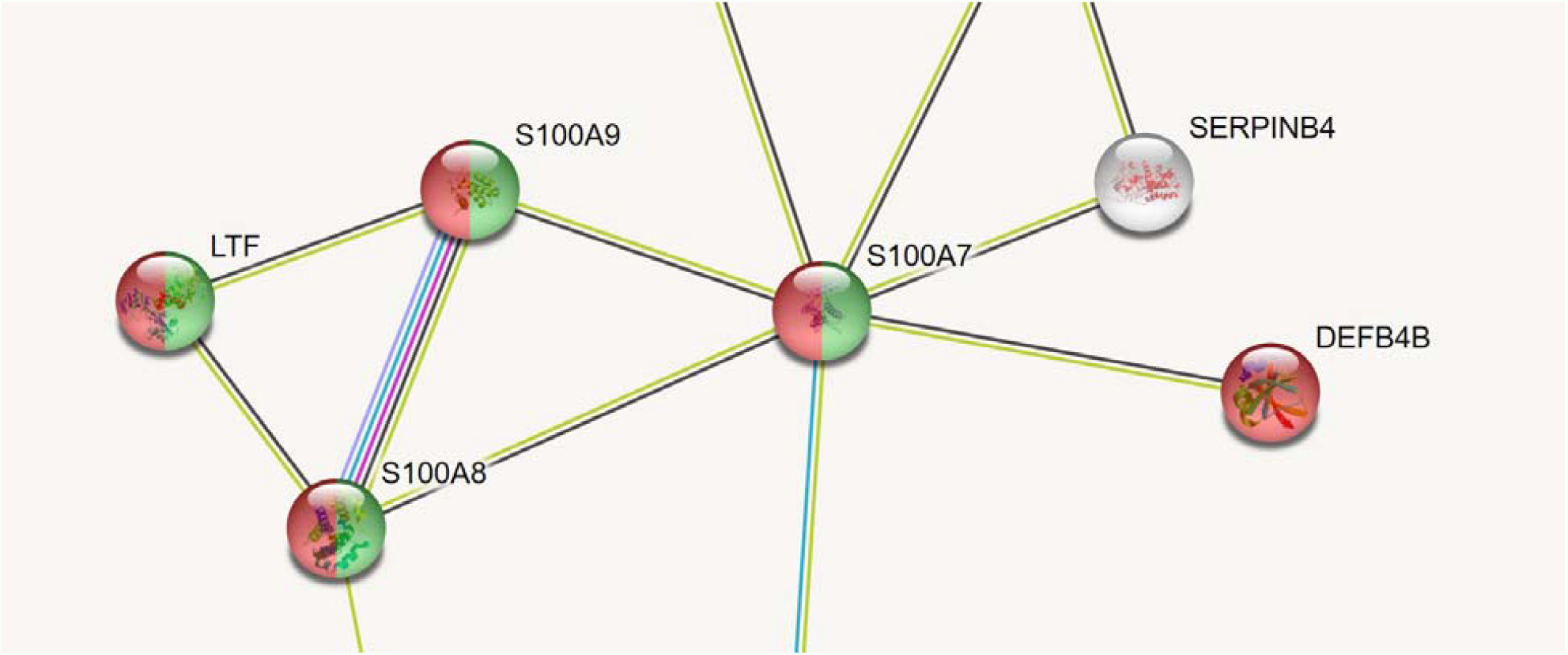
Antimicrobial peptides and metal ion sequestration gene cluster in STRING network

**Figure 4:**
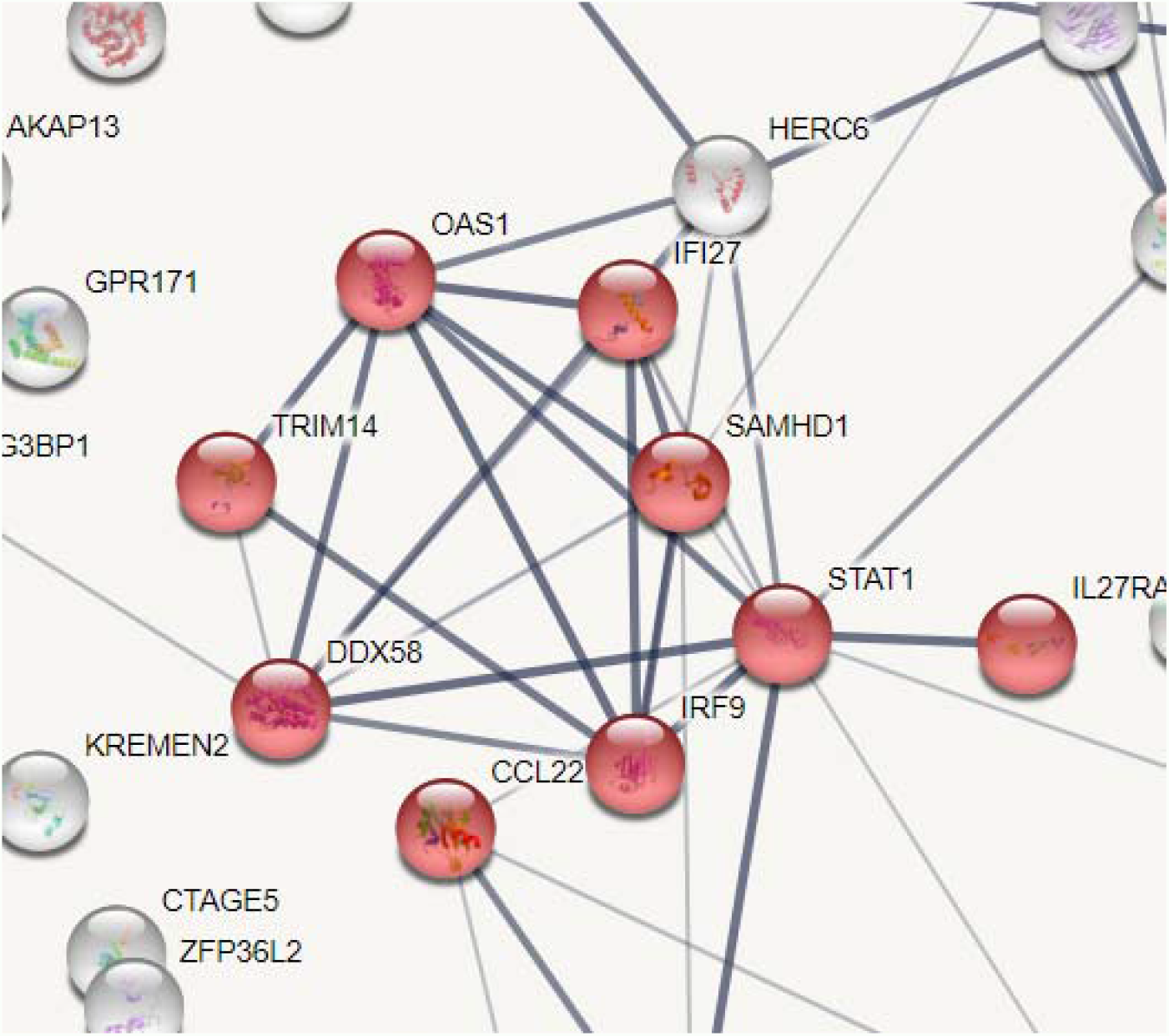
Cytokine signaling gene cluster in STRING network

Amongst upregulated genes, there was one cluster all involved in interferon signaling (red) which was not found in any of the other upregulated genes. The cluster contains STAT1, identified as a hub gene with degree 12. These genes warrant further research for exploring how interferon signaling is implicated in atopic dermatitis.

There was also a cluster of genes involved in antimicrobial peptides (red) and metal ion sequestration (green). This reflects the nature of atopic dermatitis, a disease often triggered by skin infections. Only the genes depicted were involved in antimicrobial peptides and metal ion sequestration. S100A8, S100A7, and S100A9 are also involved in RAGE receptor binding and are the only genes in the network to do so. S100A8, S100A7, DEFB4B, and S100A9 are also part of the interleukin-17 signaling pathway. SERPINB4 is shown to be connected to S100A7 and was previously identified to have the highest average fold change out of all genes. S100 proteins bind calcium which can explain the overlap between metal ion sequestration and antimicrobial peptides.

A cluster of genes involved in cytokine signaling in the immune system was prominent as well. This reflects the roots of AD in the immune system response and perhaps the inflammatory roots of AD [38-39].

Figure 5 shows all the different downregulated genes that appear in □ or in 5/5 of the datasets, and their relations to each other. 112 out of 114 genes were mapped. 68 edges were mapped with an expected value of 29 edges, again showing that there were significantly more interactions between genes than expected randomly. The average node degree was only 1.21. Genes with node degree of at least 6 were: COL1A2, LOX and DCN. All three are directly connected to each other.

**Figure 5:**
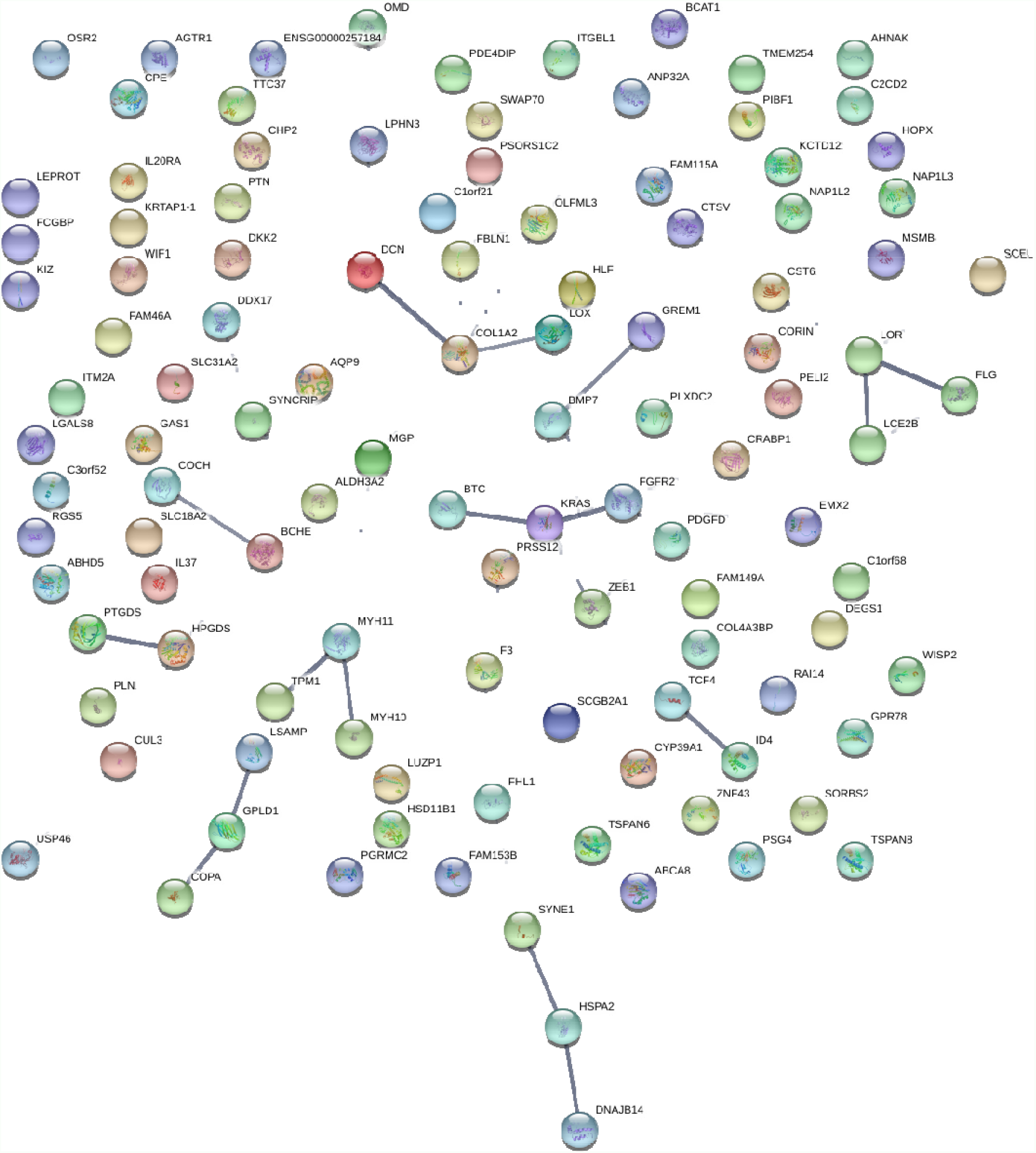
Downregulated genes network in String. Edges indicate interactions between nodes, which represent protein products of submitted genes.

The Gene Ontology component “cornified envelope” was functionally enriched. 6 genes in the GO component were found in a cluster of genes detached from the main network (Figure 6). The cornified envelope is a very tough and resistant envelope located underneath keratinocyte membranes that protects the skin [40-41]. Downregulated cornified envelope genes could partially explain skin sensitivity, disruption, and weakness in AD.

**Figure 6:**
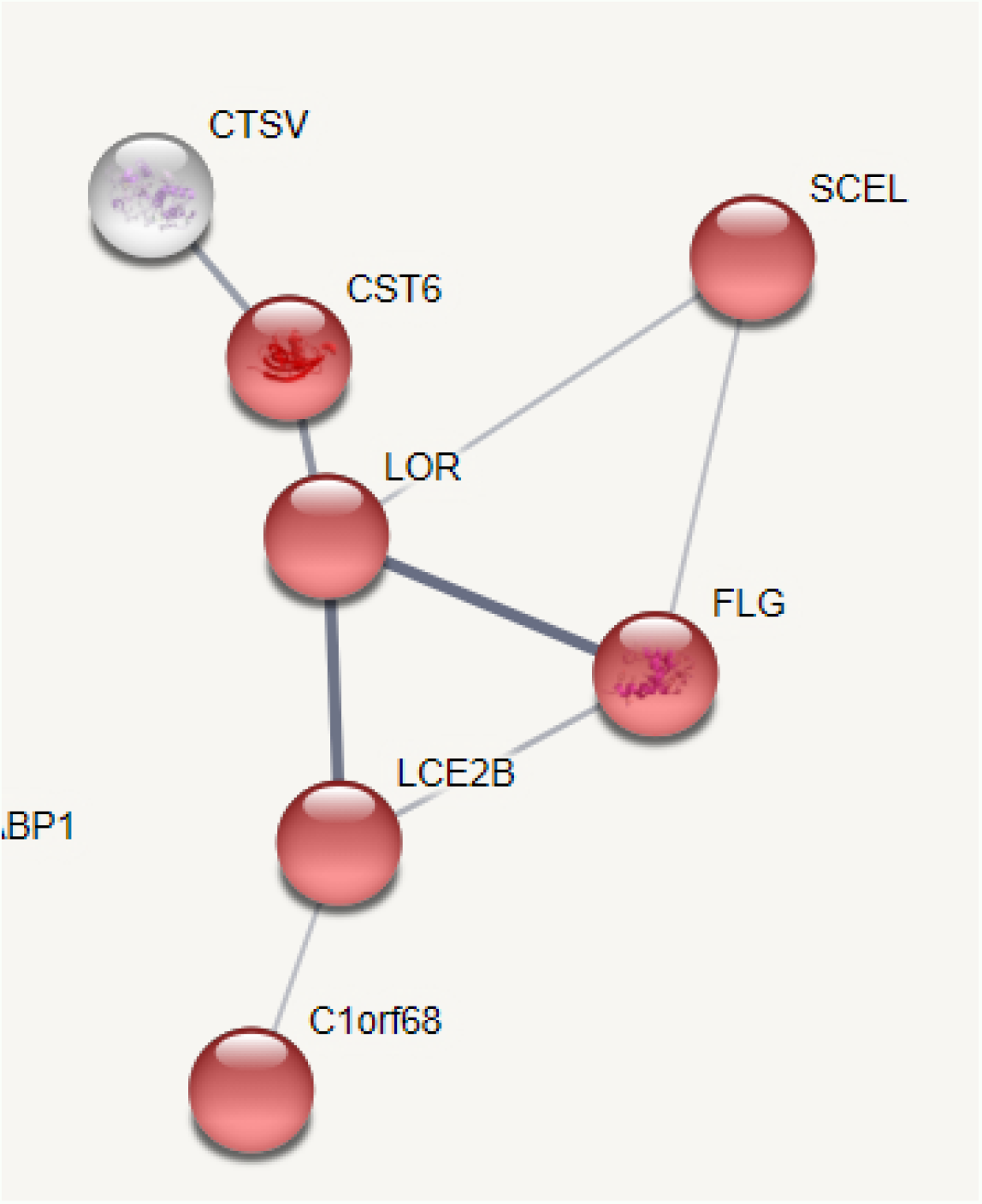
Cornified envelope cluster of genes in String network

### Panther Functional Enrichment Analysis

Upregulated and downregulated genes identified as significant in at least four of five datasets were submitted to the Gene Ontology Panther bioinformatics functional enrichment analysis tool to identify any patterns in the disrupted genes. Panther analysis revealed that the Gene Ontology Biological Process (GOBP) of neutrophil aggregation was enriched by more than 100 times (p = 1.14E-04) in the list of downregulated genes. Additionally, the GOBP Sequestering of Zinc Ion was also enriched by more than 100 times (p = 2.83E-04). The GOBPs of leukocyte aggregation, sequestering of metal ions, and type 1 interferon signaling pathway were all highly enriched as well. In total, 136 GOBPs were enriched in the downregulated genes. The first six by fold enrichment value are displayed in the table.

Amongst the upregulated gene list, there were several GOBPs identified by PantherDB however the fold enrichment was significantly less. The highest fold enrichment amongst the downregulated gene pathways was >100 while it was only 14.7 amongst the upregulated genes.

### miRNA

56 statistically significant miRNAs were identified in GSE31408 (p < 0.05). None of the overexpressed miRNAs passed both cutoff criteria (Fold Change >1.5, p < 0.05). However, 13 underexpressed miRNA passing both cutoff criteria were identified.

The miRNet software was used to predict gene targets of the 13 downregulated miRNAs [42]. The predicted gene targets were screened against the upregulated genes to identify potential interactions. For instance, a decrease in expression of a miRNA could lead to an increase in expression of its gene targets. FOXM1 was targeted by 6 out of 13 miRNAs.

## Discussion

In the present study, five gene expression datasets and one miRNA expression dataset was analyzed and commonly differentially expressed genes were identified. 208 genes were identified to be biologically relevant and found in at least C studies (94 upregulated, 114 downregulated). 14 upregulated genes and 11 downregulated genes were identified as significant in every dataset. For example, SERPINB4 had a high fold change in all datasets, including samples of chronic atopic dermatitis, nonlesional atopic dermatitis, and lesional atopic dermatitis. We also found several antimicrobial peptide genes such as S100A7 that were overexpressed. The neutrophil and leukocyte aggregation pathways were over-enriched among upregulated genes, which makes sense given the role of inflammation in AD. Cytokines and interferon signaling were also identified as over-enriched. The 25 genes, including SERPINB4, may be involved in molecular mechanisms of atopic dermatitis that are similar or the same between various subtypes of atopic dermatitis. 13 miRNAs were found to be underexpressed in AD. 6 of the 14 upregulated genes were identified as being gene targets for some of these 13 miRNAs. FOXM1 was particularly implicated, with 6 of the underexpressed miRNAs targeting it. This could explain the consistent upregulation of FOXM1 in all five datasets, with an average fold change of 9.53 meaning that FOXM1 was 9.53x more expressed in AD patients than control patients. FOXM1 is a transcription factor that is very important in many processes such as DNA repair, angiogenesis, and cell cycle progression [43]. It “regulates prolifieration, senescence, and oxidative stress in keratinocytes and cancer cells” [44]. Finally, many of the 25 consistently disrupted genes have been implicated in skin-related cancers. SERPINB4 is involved in squamous cell carcinoma [45]. FOXM1 is upregulated in basal cell carcinoma [46-47]. S100A7 is overexpressed in epithelial skin tumors [48].

The results of the study also highlight the need for caution with microarray gene expression studies. Out of thousands of statistically significant and relevantly differentially expressed genes, 63-64% of genes were only identified in a single study. This shows the great amount of variation in expression studies and perhaps serves as a warning. It could be costly and dangerous to focus research efforts on genes based on just one study. Aggregating data from multiple studies could help account for heterogeneity, methodology differences, and more, ultimately providing more reliable and dependable results. One limitation which could also be considered an advantage of the present study is that we grouped subtypes of atopic dermatitis together, for the sake of having more samples. While this means that we are able to identify common gene expression differences across subtypes, it also means that specific important gene expression differences in each subtype could be missed. Another limitation of the current study is that the datasets came from the United States and Sweden which have majority white/Caucasian populations. There could be subtle but clinically relevant racial/ethnic differences in AD that should be examined [49-50]. Overall, the present research helps elucidate molecular mechanisms of atopic dermatitis with greater confidence. Currently, corticosteroids, moisturizers, and immuno-suppressants are being used to treat AD. The identified AD-associated biological networks may be useful maps in designing new treatments for AD. The 25 consistently disrupted genes may be reliable and helpful drug targets for AD as well. Overall, this research provides insights into the molecular mechanisms behind atopic dermatitis and provides leads for treatments and diagnosis tests. The authors recommend further analysis into the 25 consistently differentially expressed genes selected in this study, as well as research into the interaction of miRNAs and mRNAs in AD disease development and progression.

## Data Availability

All data used in this study were retrieved from the public database, Gene Expression Omnibus. They are publicly available at the links below.
GSE32924: https://www.ncbi.nlm.nih.gov/geo/query/acc.cgi?acc=GSE32924
GSE6012: https://www.ncbi.nlm.nih.gov/geo/query/acc.cgi?acc=GSE6012
GSE36842: https://www.ncbi.nlm.nih.gov/geo/query/acc.cgi?acc=GSE36842
GSE16161: https://www.ncbi.nlm.nih.gov/geo/query/acc.cgi?acc=GSE16161
GSE5667: https://www.ncbi.nlm.nih.gov/geo/query/acc.cgi?acc=GSE5667
GSE31408: https://www.ncbi.nlm.nih.gov/geo/query/acc.cgi?acc=GSE31408

https://www.ncbi.nlm.nih.gov/geo/query/acc.cgi?acc=GSE6012

https://www.ncbi.nlm.nih.gov/geo/query/acc.cgi?acc=GSE36842

https://www.ncbi.nlm.nih.gov/geo/query/acc.cgi?acc=GSE16161

https://www.ncbi.nlm.nih.gov/geo/query/acc.cgi?acc=GSE5667

https://www.ncbi.nlm.nih.gov/geo/query/acc.cgi?acc=GSE31408

## Data Availability

Data used in this study were retrieved from the public database, Gene Expression Omnibus.

- GSE32924: https://www.ncbi.nlm.nih.gov/geo/query/acc.cgi?acc=GSE32924
- GSE6012: https://www.ncbi.nlm.nih.gov/geo/query/acc.cgi?acc=GSE6012
- GSE36842: https://www.ncbi.nlm.nih.gov/geo/query/acc.cgi?acc=GSE36842
- GSE16161: https://www.ncbi.nlm.nih.gov/geo/query/acc.cgi?acc=GSE16161
- GSE5667: https://www.ncbi.nlm.nih.gov/geo/query/acc.cgi?acc=GSE5667
- GSE31408: https://www.ncbi.nlm.nih.gov/geo/query/acc.cgi?acc=GSE31408

